# Simulation of low-dose PET imaging protocols for assessment of pancreatic beta-cell mass in pediatric type 1 diabetes

**DOI:** 10.64898/2026.08.17.26360614

**Authors:** Behzad Zareian, Kathryn Fontaine, Jason Bini

## Abstract

**Background:** Roughly, half of new type 1 diabetes (T1D) diagnoses occur in individuals under 18 years old and represent a more aggressive destruction of beta cell mass (BCM). [^11^C]-(+)-PHNO positron emission tomography (PET) imaging is used to assess BCM, but current pancreas PET imaging protocols are limited to adults. Previously published full count data from six healthy controls and five T1Ds (6M/5F; 22 to 53 years old) were used for retrospective analysis. Dynamic [^11^C]-(+)-PHNO PET/CT scans were acquired and reconstructed using full-count list-mode data. For the current comparison to full count data, 50%, 25% and 10% down-sampled count data were re-reconstructed. Pancreas and spleen (reference region) time-activity-curves (TACs) were assessed, and volume of distribution (*V*_T_, mL/cm^3^) was estimated using the reversible 1-tissue compartment model (1TC) with *t*_max_ of 30 min for all count levels. Pancreas and Spleen *V*_T_ estimates (1TC; *t*_max_= 30 min) were used to calculate non-displaceable binding potential (*BP*_ND_) and were then correlated to semi-quantitative methods of standardized uptake value ratio (SUVR-1) (20-30 min; ref: spleen) to examine simplified methods using simulated low dose protocols. Finally, we performed dosimetry in adult, adolescent and pediatric phantoms to assess radiation dose for simulated low-dose protocols.

**Results.:** Qualitatively, increasing noise can be visualized at successive reduced-count levels images, compared to full-count images. Despite progressively increasing noise, TACs at each reduced-count level remained similar to full-count TACs in both HC and individuals with T1D. Quantitatively, 1TC *V*_T_ estimates were similar for all reduced count levels compared to full-count (all R^2^≥0.99). Pancreas SUVR-1 (20-30 min) and pancreas *BP*_ND_ (*t*_max_ = 30; ref: spleen) were highly correlated for all count levels (all R^2^≥0.80). All age groups were under both the yearly occupational and research scan radiation dose limits when examining mean effective dose equivalent with reduced (1/10^th^) injected dose protocols.

**Conclusion.:** Low-count reconstructed data and simplified reference region approaches provide accurate quantification compared to full-count reconstructions. These results provide evidence that it is possible to perform accurate quantification using simulated low dose protocols to quantify BCM for use in individuals with T1D under 18 years old.

## Background

Type 1 diabetes (T1D) is characterized by a variable loss of pancreatic beta cell mass (BCM).[1] Roughly, half of new T1D diagnoses occur in individuals under 18 years old and represent a more aggressive and rapid destruction of BCM in pediatric populations compared to adults.[2–4]

Longitudinal magnetic resonance imaging (MRI) protocols are currently utilized to assess or monitor individuals at risk of developing T1D and during progression from stage 1 to stage 3 T1D in adolescent and pediatric populations.[5–10] Cross-sectional studies in adolescent and adult patients have been able to reveal patterns in pancreas volume, pancreatic volume index (PVI) and/or morphology metrics between matched controls and individuals with T1D increasing our understanding of disease progression. More importantly, longitudinal MRI studies have provided multiple metrics that predict progression to T1D.[7–9] However, despite revealing changes in the pancreas, MRI metrics do not provide direct assessment of BCM. Several positron emission tomography (PET) radioligands have been used to assess BCM in adult populations,[11–13] including [^11^C]-(+)-PHNO, which demonstrated utility in differentiating BCM *in vivo* between adult non-diabetic healthy controls and adult individuals with longstanding T1D.[14] However, understanding disease progression and loss of BCM using PET imaging has been, thus far, limited to adults due to radiation doses associated with PET/computed tomography (CT) scans.

The desire to combine PET BCM measurements and MRI morphology metrics, as recently shown in adults with T2D,[15] may provide a more comprehensive understanding of disease progression, when used in T1D, particularly for pediatric/adolescent onset which progresses more rapidly than adult onset T1D.[3,4] This desire motivates the development of low-dose PET acquisition protocols for pancreatic imaging. Moreover, when used in conjunction with simultaneous PET/MR scanners, replacing the CT scan with a non-ionizing radiation MRI acquisition, radiation burden on patients would be further reduced.

Recently, low-dose simulations were performed for [^11^C]-(+)-PHNO PET/MR brain imaging;[16] however, this has not been explored to date for [^11^C]-(+)-PHNO PET imaging in the pancreas. To address this need, we retrospectively analyzed our previous PET pancreas BCM imaging cohort,[14] using standard (full count) and simulated low-dose (reduced count) PET reconstructions to mimic low-dose imaging protocols. We then performed subsequent kinetic modeling to determine whether the simulated low-dose protocols provided similar quantitative accuracy in assessing BCM as the standard full count PET reconstructions. Finally, we performed full-count dosimetry in adult, adolescent and pediatric phantoms to assess radiation dose of simulated low-dose protocols.

## Materials and Methods

This is a retrospective analysis of the previously published [^11^C]-(+)-PHNO PET/CT pancreas imaging study.[14] The study was approved by the Yale University Human Investigation Committee and the Yale-New Haven Hospital Radiation Safety Committee and in accordance with federal guidelines and regulations of the USA for the protection of human research subjects contained in Title 45 Part 46 of the Code of Federal Regulations (45 CFR 46). All participants signed a written informed consent.

### PET acquisition and reconstruction

This retrospective analysis included 11 participants (age 22 to 53 years old) with six healthy controls (HC) (4M/2F) and five T1D patients (2M/3F).[14] Detailed PET acquisition methods were published previously.[14] Briefly, all participants were administered [^11^C]-(+)-PHNO simultaneously with the initiation of the PET acquisition. There were no differences in injected dose (HC: 290 ± 41 MBq, vs. T1D: 347 ± 41 MBq, p=0.05) or injected mass (HC: 0.022 ± 0.005 µg/kg, vs. T1D: 0.022 ± 0.006 µg/kg, p=0.95) between groups. [^11^C]-(+)-PHNO PET acquisitions were performed on the Siemens Biograph mCT-X PET/CT scanner with a 120-min dynamic scan (6 x 30s, 3 x 60s, 2 x 120s, and 22 x 300s) with the pancreas in the center of the field-of-view (FOV). Arterial blood sampling and metabolite analysis were performed, as described previously, to provide arterial input functions for kinetic modeling.[14] For the current study, we re-reconstructed full-count list-mode data with our in-house MOLAR PET reconstruction platform[17] using OSEM (3 iterations, 21 subsets), TOF SSS, spatially invariant PSF, all other standard corrections, and 3 mm FWHM post-recon filter and confirmed, similar to previous work,[17] that the MOLAR platform was suitable for reconstruction simulations.

### Low-dose simulation using down-sampled low-count PET data

To produce simulated low-dose PET injections, we created reduced-count PET data using previously established methods.[18,19] Each individual PET full count list-mode dataset was down-sampled by re-binning Poisson-distributed events, from the list-mode, sequentially into time-based gates. The time step of each bin was 1 second. From these sequentially time-based gates, we non-randomly selected either 50%, 25% or 10% of the count in the time-based gates. The full-count, 50%, 25% and 10% count data were then binned into the original dynamic frames (6 x 30s, 3 x 60s, 2 x 120s, and 22 x 300s) and reconstructed using the same MOLAR PET reconstruction platform and settings listed above.[17]

### Kinetic Modeling of PET data

Previously, compartmental modeling using the metabolite-corrected arterial input function and full-count [^11^C]-(+)-PHNO PET acquisitions were used to assess volume of distribution (*V*_T_, mL/cm^3^) in the pancreas and spleen and non-displaceable binding potential (*BP*_ND_) in the pancreas:[14]

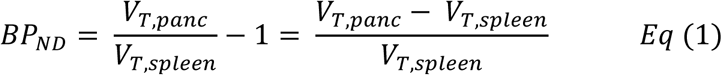

Simplified methods such as pancreas SUVR-1 (20-30 min; reference region: spleen):

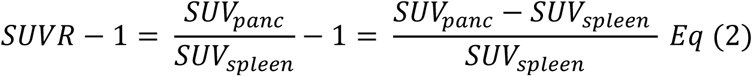

were previously validated against the gold standard compartmental modeling methods,[20] showing these methods could provide simple but accurate quantitative measures of BCM in HCs and T1Ds.[14]

Here, we performed a similar stepwise comparison from fully quantitative compartmental modeling to simplified methods using full-count and down-sampled datasets. First, we performed compartmental modeling using the metabolite-corrected arterial input function and the reversible 1-tissue compartment model (1TC), with full count data and *t*_max_ ranging from 120 min down to 30 min, to confirm stability of pancreas and spleen (reference region) *V*_T_, as previously established.[14] Subsequently, 1TC *V*_T_ estimates were determined from low count data using each respective down-sampled dataset (50%, 25%, and 10%). Pancreas and spleen *V*_T_ estimates (1TC; *t*_max_= 30 min) were then used to calculate pancreas *BP*_ND_. Linear correlations were performed between full-count and each respective low-count kinetic modeling parameter that was estimated; *K*_1_, *k*_2_, *V*_T_ and *BP*_ND_ using full-count data and *K*_1_, *k*_2_, *V*_T_ and *BP*_ND_ using 50%, 25% and 10% data. It is important to establish, not only that low-dose quantification can be accurately performed, but can be performed without arterial sampling methods, as previously established.[14] Such invasive methods are prohibitive for pediatric populations, therefore; the use of simplified protocols, removing the need for arterial sampling, must be validated. With this in mind, we performed linear correlations between pancreas *BP*_ND_ and pancreas SUVR-1 (20-30 minutes; reference: spleen) to examine whether simplified methods without the use of arterial blood sampling (SUVR-1) can be used with simulated low-dose protocols.

### Pediatric [^11^C]-(+)-PHNO Dosimetry

Patient demographics of HCs for dosimetry calculations can be found in **Table 1**. Previously published [^11^C]-(+)-PHNO dosimetry did not include a pancreas time activity curve (TAC) to calculate residence times for pancreas as input into the dosimetry calculations,[21] likely due to the fast washout of pancreas (<30 min), despite high uptake and specific binding in the pancreas.[22] In addition, dosimetry calculations for [^11^C]-(+)-PHNO using adolescent or pediatric phantoms have not been performed to date.

**Table 1.** Patient demographic information and organ sizes for dosimetry calculations.

| Diagnosis | Sex | Age (y) | Weight (kg) | Injected Dose (MBq) | Injected Mass (μg/kg) |
| --- | --- | --- | --- | --- | --- |
| HC | M | 28.7 | 81.0 | 255.2 | 0.026 |
| HC | F | 22.9 | 83.5 | 255.7 | 0.019 |
| HC | M | 26.2 | 66.5 | 318.7 | 0.027 |
| HC | M | 24.9 | 90.0 | 258.0 | 0.016 |
| HC | F | 25.7 | 65.5 | 353.3 | 0.025 |
| HC | M | 22.0 | 80.0 | 297.9 | 0.016 |

Given the pancreas and spleen are organs of interest for our pediatric studies, we calculated residence times of pancreas and spleen from our previous pancreas PET imaging cohort for input into dosimetry software, to ensure accurate organ and effective dose estimations. We also restricted dosimetry analysis to HCs to avoid underestimation of residence times in those with lower pancreas uptake (T1D), thereby avoiding skewing the results with higher allowed dose estimates in pediatric cases.

To calculate residence times, the area under the curve (AUC) was extracted from pancreas and spleen time activity curves (TACs) using the trapezoidal rule. Extrapolation from the midpoint of the last frame of the scan to infinity was performed using carbon-11 decay to determine AUC beyond the scan duration. Total AUC was then normalized to the injected dose for each patient. The resulting values were then normalized to organ masses for each standard OLINDA/EXM 1.0 model phantom (**Table 2**)[23,24] to produce residence times in hours for each organ. To use our calculated pancreas and spleen residence times in combination with previously published organ residence times,[21] each individual spleen value was added to back to the previous remainder, then pancreas and spleen residence times were subtracted to calculate an updated remainder value for input into OLINDA/EXM 1.0 software.

**Table 2.** Phantom weights and organ masses for each adult and pediatric standard OLINDA phantom.

|  | Adult Male | Adult Female | 15-year-old | 10-year-old | 5-year-old | 1-year-old |
| --- | --- | --- | --- | --- | --- | --- |
| Nominal phantom weight (kg) | 70 | 55 | 55 | 32 | 19 | 9.8 |
| Pancreas organ masses (g) | 94.3 | 85 | 64.9 | 30 | 23.6 | 10.3 |
| Spleen organ masses (g) | 183.0 | 150.0 | 123.0 | 77.4 | 48.3 | 25.5 |

For all other organs, residence times from the previous [^11^C]-(+)-PHNO dosimetry study [21] were used as input for OLINDA/EXM 1.0 software to calculate mean organ radiation doses (mSv/MBq), mean effective dose (mSv/MBq) and mean effective dose equivalent (EDE) (mSv/MBq) for each OLINDA phantom.

Yearly occupational radiation dose limits for children and adolescents under 18 are set at 10% of the adult yearly occupational limit (adult 5 rem/50 mSv; pediatric 0.5 rem/5 mSv), based on US federal regulation Section 10 CFR 20.1201.1(a)(1)(i). In combination with US federal regulation Section 21 CFR 361.1(b)(3)(ii), which sets radiation dose limits of radioactive drugs for research purposes, adults can receive either a single scan limit of 5 rem (50 mSv) or yearly organ dose limit 15 rem (150 mSv). For children and adolescents under 18, 10% of the yearly dose limit for a single scan (1.5 rem/15 mSv) is higher than the yearly occupational limit for children and adolescents (0.5 rem/5 mSv). Mean EDE measurements were used to estimate single scan radiation dose (µSv) using 1/10^th^ mean of actual injected dose in adults from our study. These single scan radiation doses were then compared to both yearly occupational (5000 µSv) and research (15000 µSv) radiation dose limits for children and adolescents under 18.

## RESULTS

Representative axial slices of fused PET/CT SUV images (20-30 min) of pancreas (green arrow) and spleen (blue arrow) can be seen in a HC using full count (**Figure 1A**), 50% (**Figure 1B**), 25% (**Figure 1C**), and 10% (**Figure 1D**) reconstructions. Representative 1TC model fits of TACs were similar for pancreas and spleen of a HC and a T1D when using full count or any of the reduced count data (**Figure 2**). For full count data, we confirmed 1TC *V*_T_ estimates for pancreas and spleen were stable when shortening from *t*_max_ of 120 min (Mean ± STD, pancreas - HC: 27.2 ± 3.1, T1D: 23.7 ± 6.6; spleen - HC: 6.2 ±1.0, T1D: 6.2 ± 1.6), to *t*_max_ of 60 min (pancreas - HC: 27.3 ± 3.5, T1D: 23.9 ± 7.0; spleen - HC: 6.1 ± 1.0, T1D: 6.2 ± 1.6) and *t*_max_ of 30 min (pancreas - HC: 28.2 ± 4.4, T1D: 24.1 ± 8.4; spleen - HC: 6.1 ± 1.0, T1D: 6.1 ± 1.6). Thus, as previously validated for vendor full count data,[14] we implemented 1TC modeling with *t*_max_ = 30 min for subsequent MOLAR low-count comparisons.

**Figure 1.**
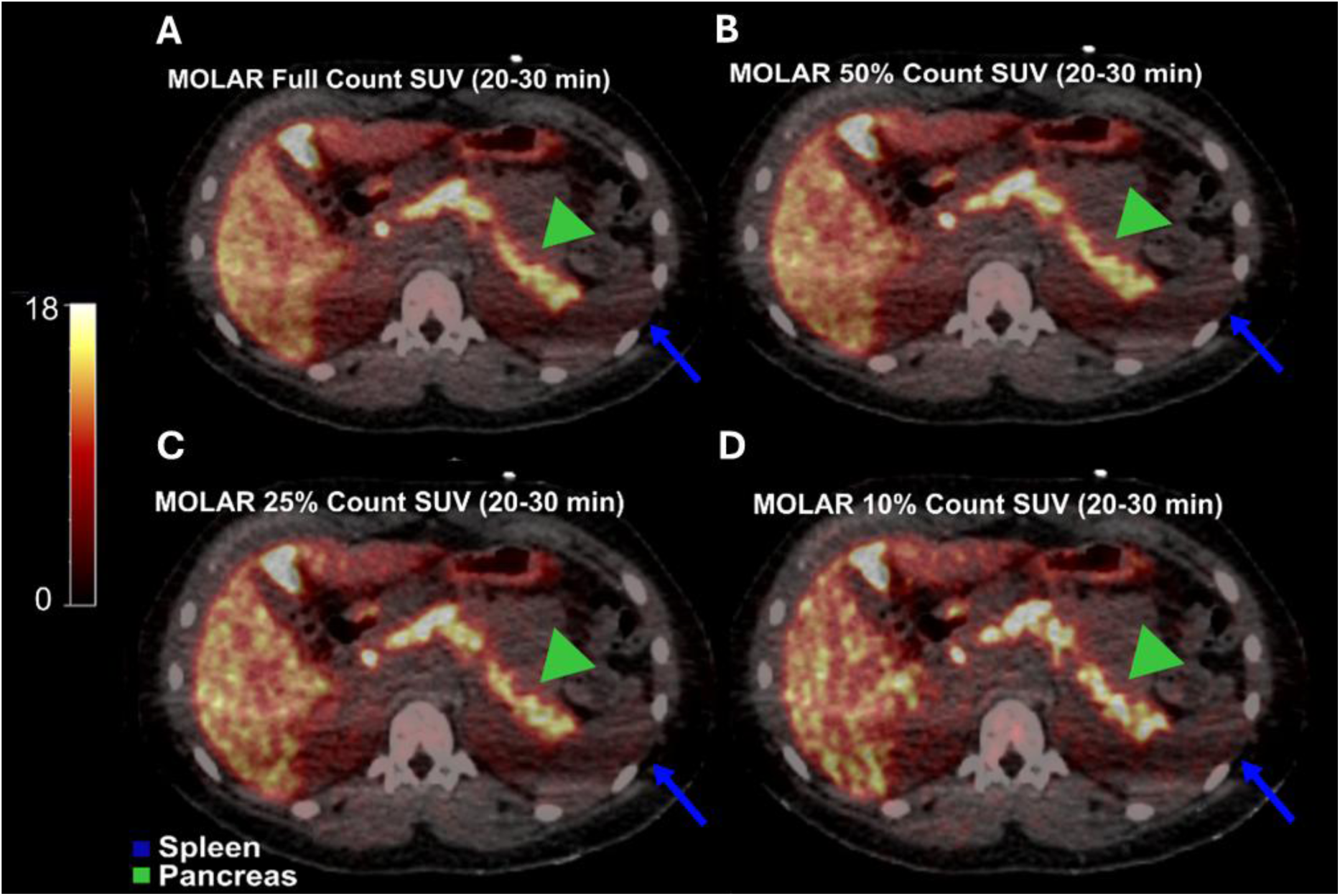
Representative axial slices of fused of [11C]-(+)-PHNO PET/CT SUV images (20-30 min) reconstructed with full (**A**), 50% (**B**), 25% (**C**), and 10% (**D**) count data. Pancreas region-of-interest (**green arrow**) and spleen reference region (**blue arrow**) can be seen in a healthy control subject. Colorbar SUV scale.

**Figure 2.**
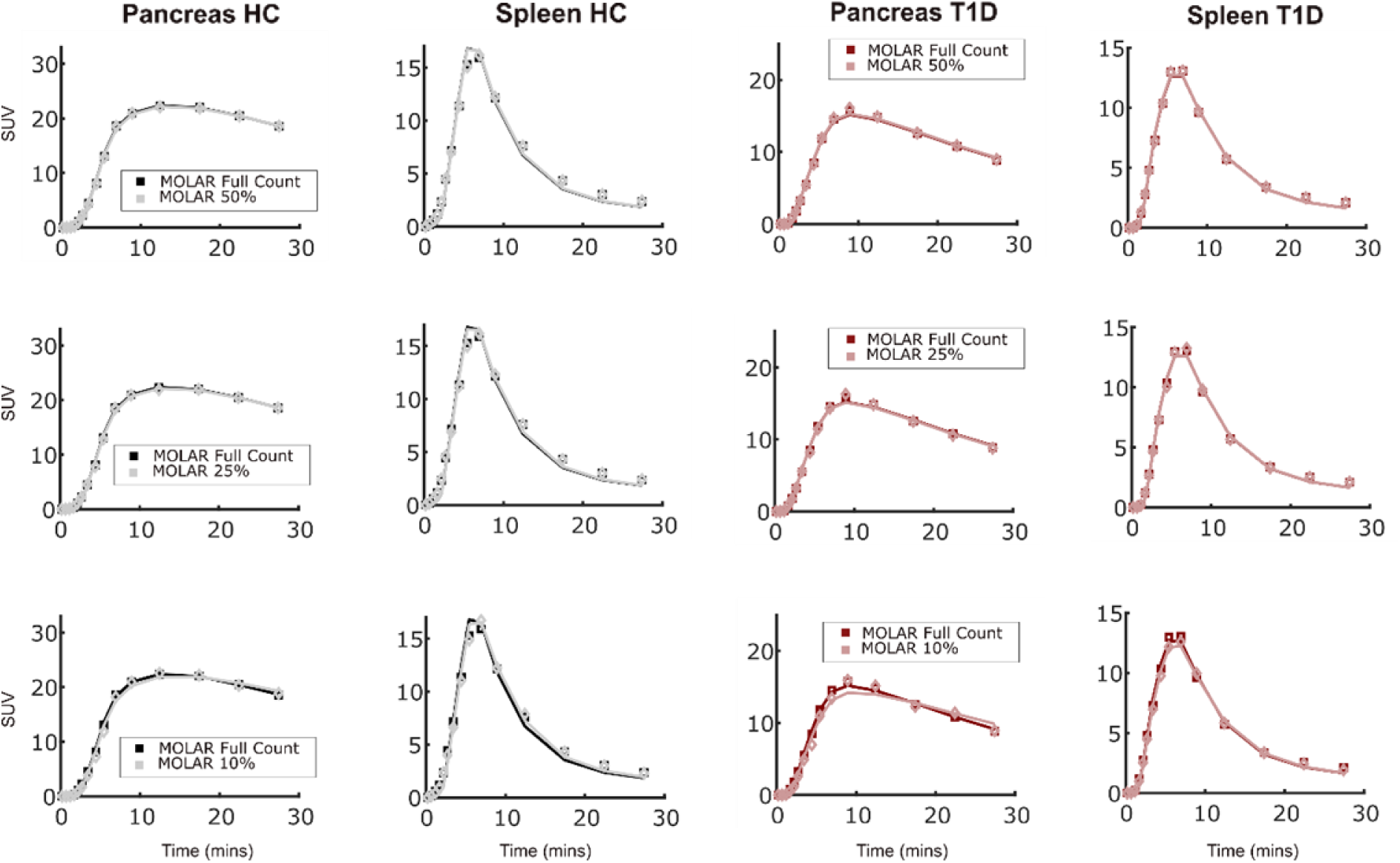
Representative time activity curves (**squares**) derived from dynamic PET frames reconstructed with full, 50%, 25% and 10% count data and 1TC model fits (**solid lines**) for pancreas and spleen in a healthy control and patient with type 1 diabetes.

In pancreas and spleen, 1TC *V*_T_ estimates for all lower count reconstructions were highly correlated to 1TC *V*_T_ estimates using full count reconstructions (all R^2^≥0.99; **Figure 3**), enabling the use of the spleen as a reference region for simplified methods in low count data to estimate *BP*_ND_ and SUVR-1 (20-30 min; ref: spleen). Rate constants *K*_1_ and *k*_2_ were highly correlated in both pancreas and spleen across all count level reconstructions (all R^2^≥0.83; **Supplemental Figure 1**). Mean *K*_1_ and *k*_2_ values and mean standard error of fit of *K*_1_ and *k*_2_ estimates were also similar for pancreas and spleen, across all count levels (**Supplemental Table 1**). *BP*_ND_ estimates remained stable across all count levels (full-count, *BP*_ND_: HC: 3.7 ± 0.8, T1D: 3.0 ± 1.0; 50% count: HC: 3.7 ± 0.8, T1D: 3.0 ± 1.0; 25% count: HC: 3.7 ± 0.8, T1D: 2.9 ± 1.0, 10% count: HC: 3.8 ± 0.8, T1D: 3.2 ± 1.0). Similarly, pancreas SUVR-1 remained stable across all count levels (full-count, SUVR-1: HC: 5.0 ± 1.1, T1D: 3.8 ± 1.2; 50% count: HC: 5.0 ± 1.1, T1D: 3.8 ± 1.1; 25% count: HC: 5.0 ± 1.1, T1D: 3.6 ± 0.9, 10% count: HC: 5.0 ± 1.2, T1D: 3.8 ± 1.1). Similar correlations were found for all count levels between SUVR-1 and *BP*_ND_ (all R^2^≥0.80; **Figure 4**). Residence times for pancreas and spleen from our current pancreas imaging protocol can be seen in **Table 3**. Mean total absorbed doses in target organs derived from the 55-kg adult female, 70-kg adult male, 15-year-old, 10-year-old, 5-year-old and 1-year old phantoms are shown in **Table 4**.

**Figure 3.**
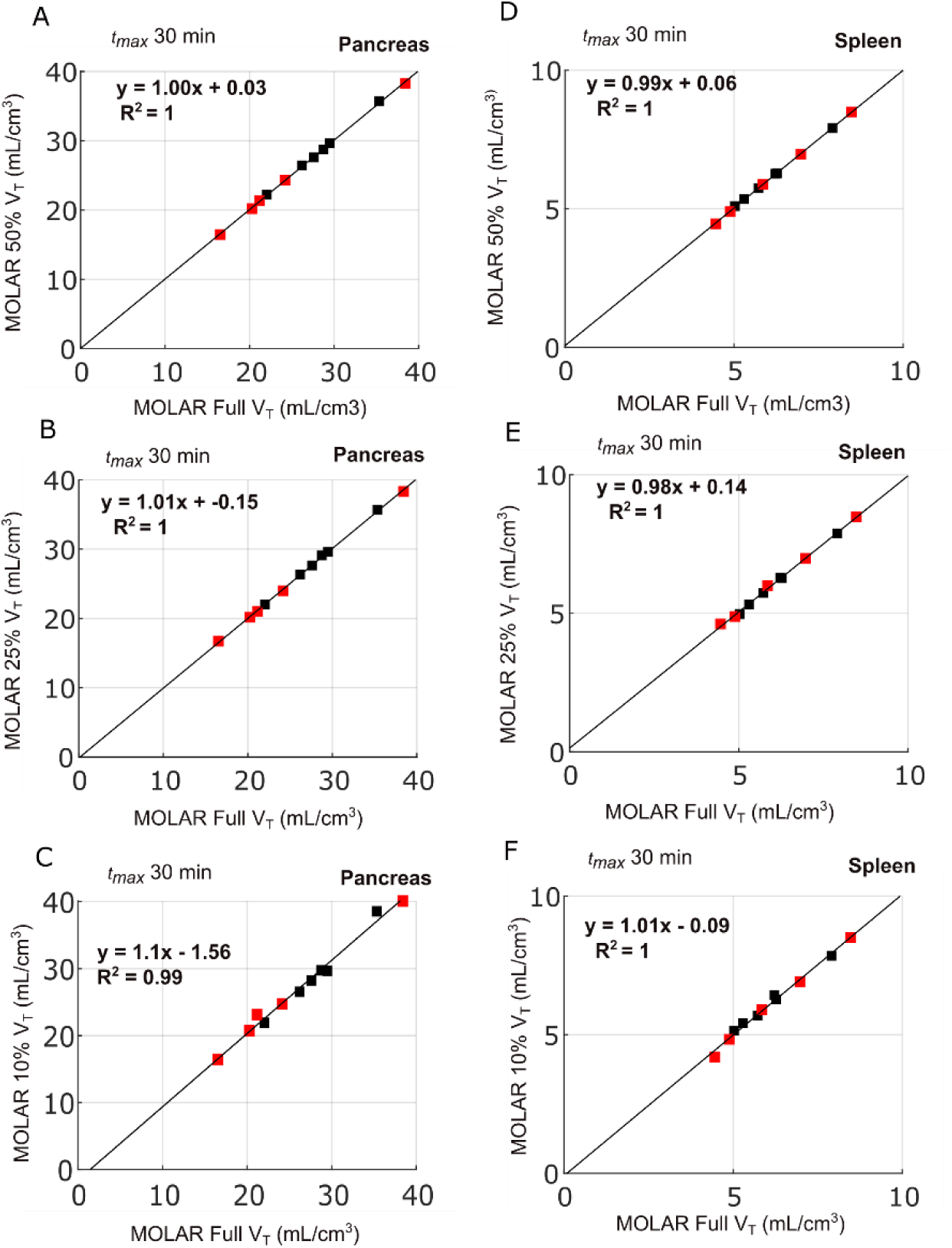
Scatter plots of volume of distribution (*VT*) estimates (1TC, *t*max=30) in pancreas (**A-C**) and spleen (**D-F**) between full-count data and reduced count data in healthy controls (**black squares**) and individuals with type 1 diabetes (**red squares**). Black line represents linear regression fit.

**Figure 4.**
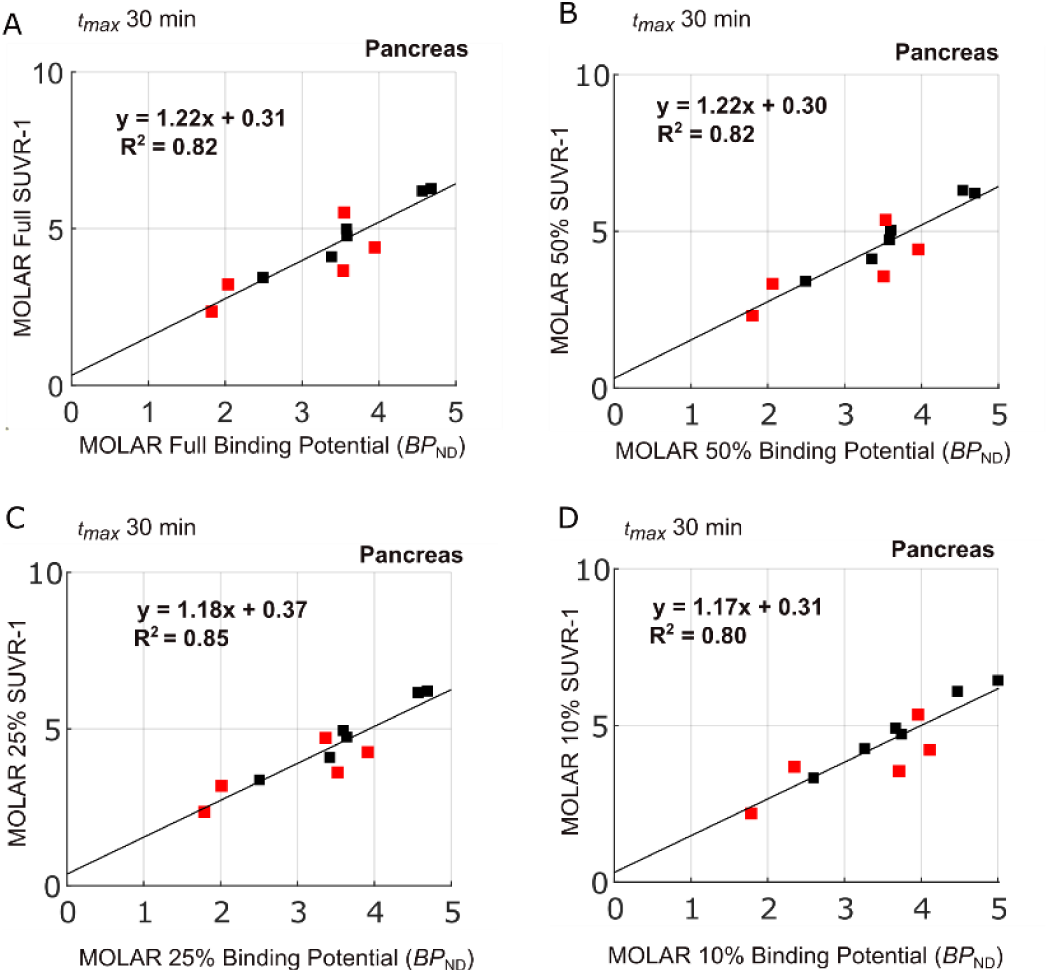
Scatter plots of non-displaceable binding potential (*BPND*) estimates and SUVR-1 (20-30 min) in pancreas between full-count data (**A**) and 50% reduced count data (**B**) 25% reduced count data (**C**) and 10% reduced count data (**D**) in healthy controls (**black squares**) and individuals with type 1 diabetes (**red squares**). Black line represents linear regression fit.

**Table 3.** Residence Times (hours). Bold values for pancreas and spleen are calculated from our current data. The remainder is also altered to reflect the addition of the calculated pancreas residence time. All other organ residence times were used from the previous dosimetry study (**Mizrahi, 2012**) as input in OLINDA/EXM 1.0 software.

| Organ | Females | Males | 15-year-old | 10-year-old | 5-year-old | 1-year-old |
| --- | --- | --- | --- | --- | --- | --- |
| Brain | 0.029 ± 0.005 | 0.027 ± 0.007 | 0.029 ± 0.005 | 0.029 ± 0.005 | 0.029 ± 0.005 | 0.029 ± 0.005 |
| Lung | 0.027 ± 0.007 | 0.027 ± 0.009 | 0.027 ± 0.007 | 0.027 ± 0.007 | 0.027 ± 0.007 | 0.027 ± 0.007 |
| Liver | 0.073 ± 0.011 | 0.102 ± 0.024 | 0.073 ± 0.011 | 0.073 ± 0.011 | 0.073 ± 0.011 | 0.073 ± 0.011 |
| Pancreas | **0.0084 ± 0.0001** | **0.0092 ± 0.0004** | **0.0075 ± 0.0009** | **0.006 ± 0.0007** | **0.0079 ± 0.0010** | **0.0067 ± 0.0008** |
| Spleen | **0.0051 ± 0.0003** | **0.0058 ± 0.001** | **0.0047 ± 0.0008** | **0.0051 ± 0.0009** | **0.0053 ± 0.0009** | **0.0055 ± 0.0009** |
| Kidneys | 0.014 ± 0.004 | 0.014 ± 0.004 | 0.014 ± 0.004 | 0.014 ± 0.004 | 0.014 ± 0.004 | 0.014 ± 0.004 |
| Bladder | 0.011 ± 0.005 | 0.017 ± 0.005 | 0.017 ± 0.005 | 0.017 ± 0.005 | 0.017 ± 0.005 | 0.017 ± 0.005 |
| Remainder | **0.3225 ± 0.0005** | **0.288 ± 0.0013** | **0.3238 ± 0.0015** | **0.325 ± 0.0015** | **0.3227 ± 0.0017** | **0.3238 ± 0.0016** |

**Table 4.** Mean ± standard deviation (SD) of organ radiation doses (mSv/MBq), effective dose equivalent (mSv/MBq) and effective dose (mSv/MBq). Respective OLINDA phantoms weights (kg).

| (mSv/MBq) | **Adult female (55 kg)** |  | **Adult male (70 kg)** |  | **15-year-old (55 kg)** |  |
| --- | --- | --- | --- | --- | --- | --- |
| **Organ** | **Mean** | **± SD** | **Mean** | **± SD** | **Mean** | **± SD** |
| Adrenals | 4.52E-03 | 7.07E-06 | 3.72E-03 | 9.57E-06 | 4.46E-03 | 2.59E-05 |
| Brain | 8.07E-03 | 0.00E+00 | 6.46E-03 | 0.00E+00 | 7.04E-03 | 4.08E-06 |
| Breasts | 2.56E-03 | 7.07E-06 | 1.92E-03 | 5.77E-06 | 2.56E-03 | 8.16E-06 |
| Gallbladder Wall | 4.88E-03 | 0.00E+00 | 4.50E-03 | 5.77E-06 | 4.80E-03 | 1.47E-05 |
| LLI Wall | 3.15E-03 | 0.00E+00 | 2.35E-03 | 5.00E-06 | 2.97E-03 | 1.05E-05 |
| Small Intestine | 3.18E-03 | 7.07E-06 | 2.58E-03 | 5.77E-06 | 3.40E-03 | 7.53E-06 |
| Stomach Wall | 3.71E-03 | 1.41E-05 | 2.89E-03 | 1.91E-05 | 3.60E-03 | 3.45E-05 |
| ULI Wall | 3.47E-03 | 0.00E+00 | 2.69E-03 | 5.00E-06 | 3.40E-03 | 5.16E-06 |
| Heart Wall | 3.75E-03 | 0.00E+00 | 2.88E-03 | 0.00E+00 | 3.78E-03 | 4.08E-06 |
| Kidneys | 1.55E-02 | 0.00E+00 | 1.42E-02 | 0.00E+00 | 1.69E-02 | 0.00E+00 |
| Liver | 1.74E-02 | 0.00E+00 | 1.80E-02 | 0.00E+00 | 1.76E-02 | 5.48E-05 |
| Lungs | 1.02E-02 | 0.00E+00 | 8.15E-03 | 0.00E+00 | 1.19E-02 | 0.00E+00 |
| Muscle | 2.84E-03 | 7.07E-06 | 2.15E-03 | 5.00E-06 | 2.85E-03 | 7.53E-06 |
| Ovaries | 3.22E-03 | 7.07E-06 | 2.45E-03 | 9.57E-06 | 3.28E-03 | 1.17E-05 |
| Pancreas | **2.76E-02** | **4.24E-04** | **2.70E-02** | **1.04E-03** | **3.16E-02** | **3.48E-03** |
| Red Marrow | 2.70E-03 | 0.00E+00 | 2.12E-03 | 0.00E+00 | 2.77E-03 | 5.16E-06 |
| Osteogenic Cells | 4.29E-03 | 7.07E-06 | 2.97E-03 | 9.57E-06 | 4.25E-03 | 1.41E-05 |
| Skin | 2.25E-03 | 0.00E+00 | 1.67E-03 | 8.16E-06 | 2.24E-03 | 8.94E-06 |
| Spleen | 1.13E-02 | 6.36E-04 | 1.02E-02 | 1.57E-03 | 1.21E-02 | 1.76E-03 |
| Testes |  |  |  | 1.91E-03 | 8.16E-06 | 2.62E-03 |
| Thymus | 3.00E-03 | 7.07E-06 | 2.19E-03 | 5.77E-06 | 3.03E-03 | 1.05E-05 |
| Thyroid | 2.57E-03 | 7.07E-06 | 2.00E-03 | 5.77E-06 | 2.82E-03 | 1.05E-05 |
| Urinary Bladder | 1.25E-02 | 0.00E+00 | 1.34E-02 | 0.00E+00 | 1.21E-02 | 0.00E+00 |
| Uterus | 3.38E-03 | 0.00E+00 | 2.73E-03 | 9.57E-06 | 3.48E-03 | 1.17E-05 |
| Total Body | 3.62E-03 | 7.07E-06 | 2.87E-03 | 5.00E-06 | 3.62E-03 | 5.16E-06 |
| Effective Dose Equivalent (mSv/MBq) | 8.00E-03 | 6.36E-05 | 7.25E-03 | 1.38E-04 | 8.60E-03 | 2.83E-04 |
| Effective Dose (mSV/MBq) | 5.67E-03 | 7.07E-06 | 4.96E-03 | 2.52E-05 | 5.96E-03 | 9.16E-05 |

| (mSv/MBq) | 10-year-old (32 kg) |  | 5-year-old (19 kg) |  | 1-year-old (9.8 kg) |  |
| --- | --- | --- | --- | --- | --- | --- |
| Organ | Mean | ± SD | Mean | ± SD | Mean | ± SD |
| Adrenals | 6.86E-03 | 3.33E-05 | 1.07E-02 | 5.16E-05 | 1.90E-02 | 6.32E-05 |
| Brain | 7.45E-03 | 5.16E-06 | 8.39E-03 | 4.08E-06 | 1.17E-02 | 0.00E+00 |
| Breasts | 4.11E-03 | 1.17E-05 | 6.58E-03 | 1.87E-05 | 1.27E-02 | 5.16E-05 |
| Gallbladder Wall | 7.13E-03 | 4.04E-05 | 1.13E-02 | 5.16E-05 | 2.08E-02 | 9.83E-05 |
| LLI Wall | 4.92E-03 | 1.72E-05 | 7.80E-03 | 2.56E-05 | 1.44E-02 | 7.53E-05 |
| Small Intestine | 5.50E-03 | 1.17E-05 | 8.84E-03 | 1.86E-05 | 1.67E-02 | 5.16E-05 |
| Stomach Wall | 5.59E-03 | 4.17E-05 | 9.05E-03 | 6.77E-05 | 1.69E-02 | 8.94E-05 |
| ULI Wall | 5.53E-03 | 8.37E-06 | 8.91E-03 | 1.51E-05 | 1.68E-02 | 5.16E-05 |
| Heart Wall | 5.91E-03 | 4.08E-06 | 9.13E-03 | 7.53E-06 | 1.68E-02 | 0.00E+00 |
| Kidneys | 2.38E-02 | 5.16E-05 | 3.55E-02 | 8.16E-05 | 6.31E-02 | 1.03E-04 |
| Liver | 2.64E-02 | 3.80E-18 | 3.92E-02 | 7.60E-18 | 7.41E-02 | 5.16E-05 |
| Lungs | 1.69E-02 | 4.08E-05 | 2.60E-02 | 0.00E+00 | 5.12E-02 | 4.08E-05 |
| Muscle | 4.53E-03 | 8.16E-06 | 7.24E-03 | 1.72E-05 | 1.40E-02 | 5.16E-05 |
| Ovaries | 5.24E-03 | 1.72E-05 | 8.37E-03 | 3.01E-05 | 1.59E-02 | 6.32E-05 |
| **Pancreas** | **5.25E-02** | **5.99E-03** | **8.73E-02** | **9.76E-03** | **1.64E-01** | **1.86E-02** |
| Red Marrow | 4.22E-03 | 8.94E-06 | 6.73E-03 | 2.16E-05 | 1.65E-02 | 6.32E-05 |
| Osteogenic Cells | 6.07E-03 | 1.87E-05 | 9.27E-03 | 3.29E-05 | 1.95E-02 | 7.53E-05 |
| Skin | 3.62E-03 | 1.05E-05 | 5.89E-03 | 2.28E-05 | 1.15E-02 | 5.16E-05 |
| Spleen | 1.97E-02 | 2.99E-03 | 3.26E-02 | 4.76E-03 | 6.09E-02 | 8.99E-03 |
| Testes | 4.22E-03 | 1.63E-05 | 6.70E-03 | 3.01E-05 | 1.32E-02 | 7.53E-05 |
| Thymus | 4.72E-03 | 1.72E-05 | 7.44E-03 | 2.56E-05 | 1.42E-02 | 5.16E-05 |
| Thyroid | 4.53E-03 | 1.63E-05 | 7.43E-03 | 3.31E-05 | 1.42E-02 | 6.32E-05 |
| Urinary Bladder | 1.86E-02 | 3.80E-18 | 2.93E-02 | 5.16E-05 | 5.50E-02 | 5.16E-05 |
| Uterus | 5.55E-03 | 1.63E-05 | 8.88E-03 | 3.01E-05 | 1.67E-02 | 6.32E-05 |
| Total Body | 5.78E-03 | 5.16E-06 | 9.28E-03 | 5.16E-06 | 1.78E-02 | 0.00E+00 |
| Effective Dose Equivalent (mSv/MBq) | 1.32E-02 | 4.72E-04 | 2.09E-02 | 7.94E-04 | 4.00E-02 | 1.51E-03 |
| Effective Dose (mSv/MBq) | 9.15E-03 | 1.53E-04 | 1.42E-02 | 4.31E-04 | 2.74E-02 | 8.52E-04 |

The dose-limiting organ was the pancreas for both adult males (0.027 mSv/MBq) and for adult females (0.028 mSv/MBq) with yearly single-study dose limits of 1853 MBq (50.1 mCi) and 1796 MBq (48.5 mCi), respectively. The dose-limiting organ in the 15-year-old, 10-year-old, 5-year-old and 1-year old phantoms was also the pancreas with 0.032 mSv/MBq, 0.053 mSv/MBq, 0.087 mSv/MBq, and 0.164 mSv/MBq, respectively. Based on US federal regulations recommending 1/10^th^ of the adult dose, the yearly single-study radiation dose limit for pediatric scans is 158.2 MBq (4.3 mCi) for 15-year-old, 95.3 MBq (2.6 mCi) for 10-year-old, 57.3 MBq (1.5 mCi) for 5-year-old, and 30.5 MBq (0.8 mCi) for 1-year old. The mean effective dose (ED) was 4.96 μSv/MBq for adult males 5.67 μSv/MBq for adult females. The mean ED using the 15-year-old, 10-year-old, 5-year-old and 1-year old phantoms were 5.96 μSv/MBq, 9.15 μSv/MBq, 14.2 μSv/MBq, and 27.4 μSv/MBq, respectively.

All age groups, including adults, are under both the yearly occupational and research scan radiation dose limits when examining mean EDE and reduced (1/10^th^) injected dose protocols (**Table 5**). Using 1/10^th^ of the actual injected dose from our healthy control participants, all ages from 10-years-old and greater would receive ≤8% of the yearly occupational radiation dose limit or ≤3% of the yearly research participant dose limit (**Table 5**). Occupational yearly limits for 5-year-olds and 1-year-olds were higher at 12% and 23%, respectively. Research participant yearly limits for 5-year-olds and 1-year-olds were also slightly higher, at 4% and 8%, respectively.

**Table 5.**
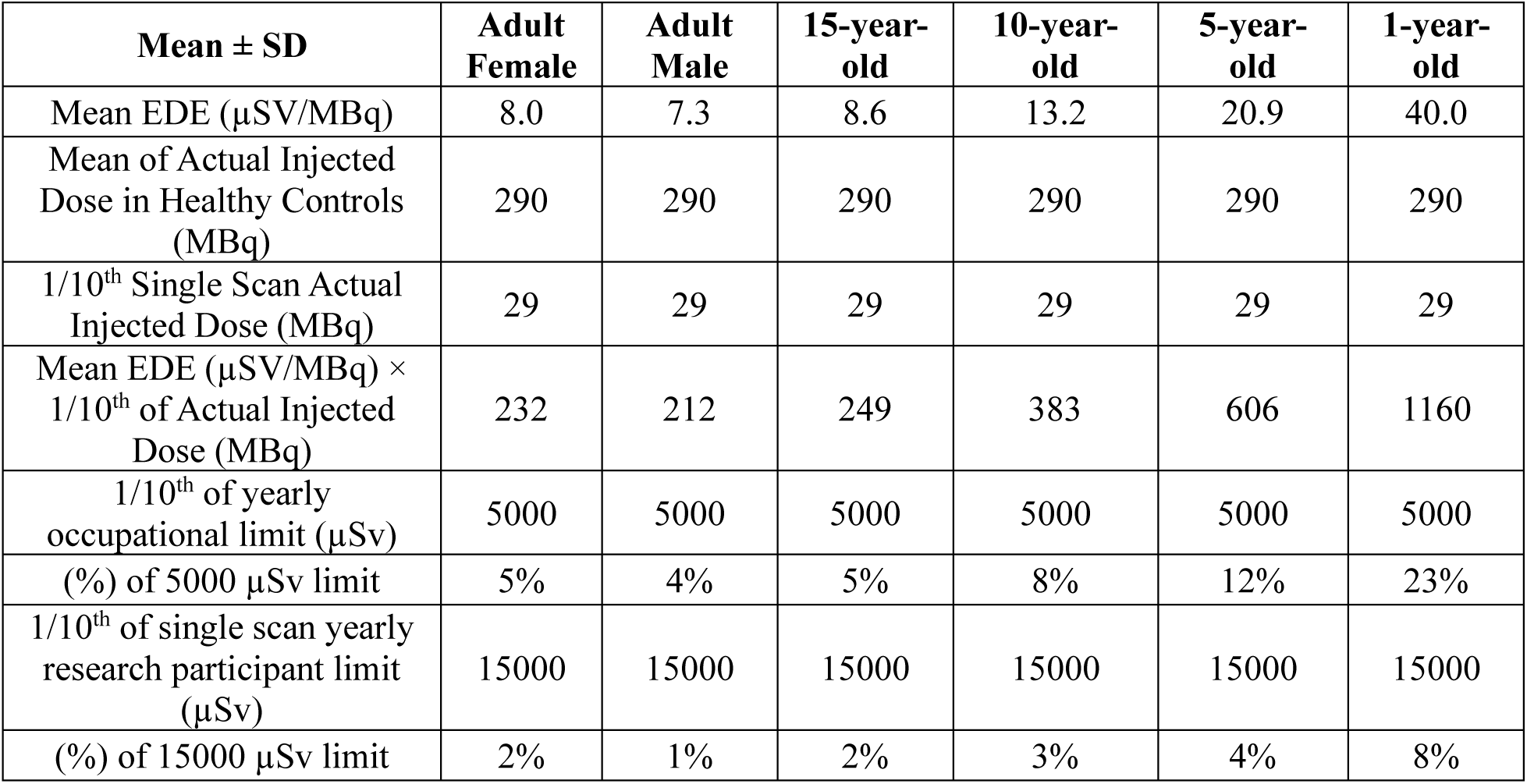
Mean Effective Dose Equivalent measurements from Olinda dosimetry software with 1/10^th^ actual injected dose in healthy controls and comparison to yearly occupational and research radiation dose limit for children and adolescents under 18 years old.

| Mean ± SD | Adult Female | Adult Male | 15-year-old | 10-year-old | 5-year-old | 1-year-old |
| --- | --- | --- | --- | --- | --- | --- |
| Mean EDE ($\mu\text{SV}/\text{MBq}$) | 8.0 | 7.3 | 8.6 | 13.2 | 20.9 | 40.0 |
| Mean of Actual Injected Dose in Healthy Controls (MBq) | 290 | 290 | 290 | 290 | 290 | 290 |
| $1/10^{\text{th}}$ Single Scan Actual Injected Dose (MBq) | 29 | 29 | 29 | 29 | 29 | 29 |
| Mean EDE ($\mu\text{SV}/\text{MBq}$) × $1/10^{\text{th}}$ of Actual Injected Dose (MBq) | 232 | 212 | 249 | 383 | 606 | 1160 |
| $1/10^{\text{th}}$ of yearly occupational limit ($\mu\text{Sv}$) | 5000 | 5000 | 5000 | 5000 | 5000 | 5000 |
| (%) of 5000 $\mu\text{Sv}$ limit | 5% | 4% | 5% | 8% | 12% | 23% |
| $1/10^{\text{th}}$ of single scan yearly research participant limit ($\mu\text{Sv}$) | 15000 | 15000 | 15000 | 15000 | 15000 | 15000 |
| (%) of 15000 $\mu\text{Sv}$ limit | 2% | 1% | 2% | 3% | 4% | 8% |

## DISCUSSION

In this study, we retrospectively analyzed our previous PET pancreas BCM imaging cohort,[14] using standard (full count) and simulated low-dose (reduced count) PET reconstructions to mimic low-dose imaging protocols. Subsequent compartmental modeling and simplified methods determined that simulated low-dose protocols provided similar quantitative accuracy in assessing BCM as the standard full count PET reconstructions. Finally, we estiamted dosimetry in adult, adolescent and pediatric phantoms to assess radiation dose of simulated low-dose protocols and demonstrated that radiation dose of simulated low-dose protocols was well below the recommended yearly limit for adolescents and children under 18 years of age.

Qualitatively, increasing noise can be visualized at successive reduced-count levels in reconstructions (**Figure 1B-D**), compared to full-count reconstruction (**Figure 1A**), particularly evident in the liver. Despite the progressively increased noise in each successive reduced-count reconstructed image, TACs at each reduced-count level remained similar to full-count TACs in both HC and individuals with T1D (**Figure 2**). Compartmental modeling (1TC) TAC fits were also similar for all reduced count levels (**Figure 2**). *V*_T_ estimates using MOLAR remained stable for the range of *t*_max_ values (30-120 min) for full-count data, similar to previous vendor full-count reconstructions.[14] This indicates that we can reliably estimate *V*_T_ in both target (pancreas) and reference region (spleen) using the shortest *t*_max_ (30 min) and the lowest count level (10%) (**Figure 3**), allowing for shorter PET acquisitions to maximize patient comfort. To further reduce patient burden, it is also important to investigate simplified reference region approaches (*e.g.,* SUVR-1). *BP*_ND_ and SUVR-1 are both reference region approaches; however, *BP*_ND_ relies on estimating *V*_T_ for both target and reference region using gold standard kinetic modeling methods which require a metabolite-corrected arterial input function (**Equation 1**), whereas SUVR-1 removes the need for arterial sampling and metabolite correction by using pancreas and spleen SUV as simplified measures (**Equation 2**). In our study, SUVR-1 (20-30 min; spleen reference) was highly correlated with *BP*_ND_ at all reduced-count levels (R^2^≥0.80) and can be used as a surrogate measure of specific binding to assess BCM. Therefore, for reduced count data simulating a 10% injected dose, a short 30 min scan with no arterial sampling necessary can reliably be performed using the simplified quantitative measure SUVR-1 (20-30 min; spleen reference), as was established previously with full-count data.[14]

To examine radiation dosimetry of the proposed 10% dose, we re-examined previous [^11^C]-(+)-PHNO PET dosimetry.[21] Due to the high uptake of the pancreas, not previously included in the residence times used to calculate organ doses, we used our current cohort of healthy controls to include our pancreas and spleen data in conjunction with the previously published residence times.[21] Importantly, when re-calculating the organ doses (**Table 4**), the pancreas was the dose limiting organ when using each OLINDA phantom, rather than the liver, as established previously.[21] Despite this, the ED for both studies remains similar, although slightly higher, in adults in our study (F: 5.7 µSv/MBq; M: 5.0 µSv/MBq), compared to the previous [^11^C]-(+)-PHNO adult dosimetry (F: 5.2 µSv/MBq; M: 4.5 µSv/MBq).[21]

Finally, we calculated the estimated dose (µSv) for a single low-dose (10%) scan protocol across the range of OLINDA phantoms (**Table 5**). When using 1/10^th^ of the actual injected dose of our healthy control cohort to calculate single scan dose limits, we found that all ages from 10-years-old and greater would receive ≤8% of the yearly occupational radiation dose limit (5,000 µSv) or ≤3% of the yearly research participant dose limit (15,000 µSv)(**Table 5**). The occupational dose limit is more conservative than the research scan limit and in examining the yearly occupational limits for 5-year-olds and 1-year-olds these were higher at 12% and 23%, respectively. Taking into consideration typical difficulties in imaging young children and infants (*e.g.*, movement during PET or MRI scans), it would seem reasonable to apply low-dose [^11^C]-(+)-PHNO PET acquisition protocols in T1D patients older than 10 years of age, where these issues may be less prevalent. One caveat in the current study is that these acquisitions are performed on PET/CT scanners and thus the radiation from the low-dose CT attenuation scan is still the limiting dose. The use of simultaneous PET/MR scanners will further reduce the radiation burden and allow for acquisition of pancreas volume and shape metrics in conjunction with PET BCM.[15]

In a similar manner, Ribeiro, *et al.*, explored simulated low activity [^11^C]-(+)-PHNO datasets; however, in pursuit of low-dose protocols for neuropsychiatric imaging applications in adolescents.[16] With the benefit of acquisitions on a PET/MR scanner eliminating CT-dose, they were able to demonstrate similar quantitative estimations of *BP*_ND_ down to a simulated 1/6^th^ of the injected dose. Below this, at 1/10^th^ and 1/16^th^ of the injected dose, the coefficient of variation in the substantia nigra increased, creating unreliable *BP*_ND_ estimates. The substantia nigra, a much smaller region than the pancreas, is more susceptible to increased noise with activity reduction and not surprising compared to our results.

Our current study explored down to 10% of injected dose and our quantitative parameters estimates were similar to full count data. It may be possible that further dose reduction in combination with deep learning techniques to reduce image noise and improved quantitative accuracy of low count data could be employed.[25] Others have employed deep learning-based denoising to explore reduced scan time for pediatric imaging.[26] Given that our simplified measure is SUVR-1 (20-30min), it is already possible to perform acquisitions by injecting the adult patients, allowing a 20-minute radiotracer circulation time and imaging from 20-30 minutes post injection, conducive to pediatric imaging. Further reduction of scan times remains to be validated on simultaneous PET/MR scanners.

This validation of a low-dose [^11^C]-(+)-PHNO PET protocol, when employed on simultaneous PET/MR scanners, will also allow increased throughput. Rather than separate pancreas PET and MRI acquisitions, we can perform these studies simultaneously which is advantageous for not only adult studies, but more so in adolescent and pediatric imaging.

## Conclusion

Low count reconstructed data and simplified reference region approaches provide highly similar quantification to full-count reconstructions. These results provide evidence that it is possible to perform accurate quantification using simulated low dose protocols to quantify BCM for use in individuals with T1D under 18 years old. These protocols may allow for PET imaging of more aggressive and rapid progressive T1D, expanding our knowledge of *in vivo* BCM loss in adolescent and pediatric populations.

## Availability of data and material

The datasets used and/or analyzed during the current study are available from the corresponding author on reasonable request.

## Authors’ contributions

BZ: Investigation, Formal analysis, Methodology, Writing – original draft, Writing – review & editing, KF: Conceptualization, Writing – review & editing, Methodology, Formal analysis, Investigation. JB: Funding acquisition, Supervision, Project administration, Methodology, Conceptualization, Investigation, Formal analysis, Writing – original draft, Writing – review & editing.

## Acknowledgements

The authors appreciate the excellent technical assistance of the Yale PET Center staff.

## Supplemental Data

**Supplemental Figure 1.**
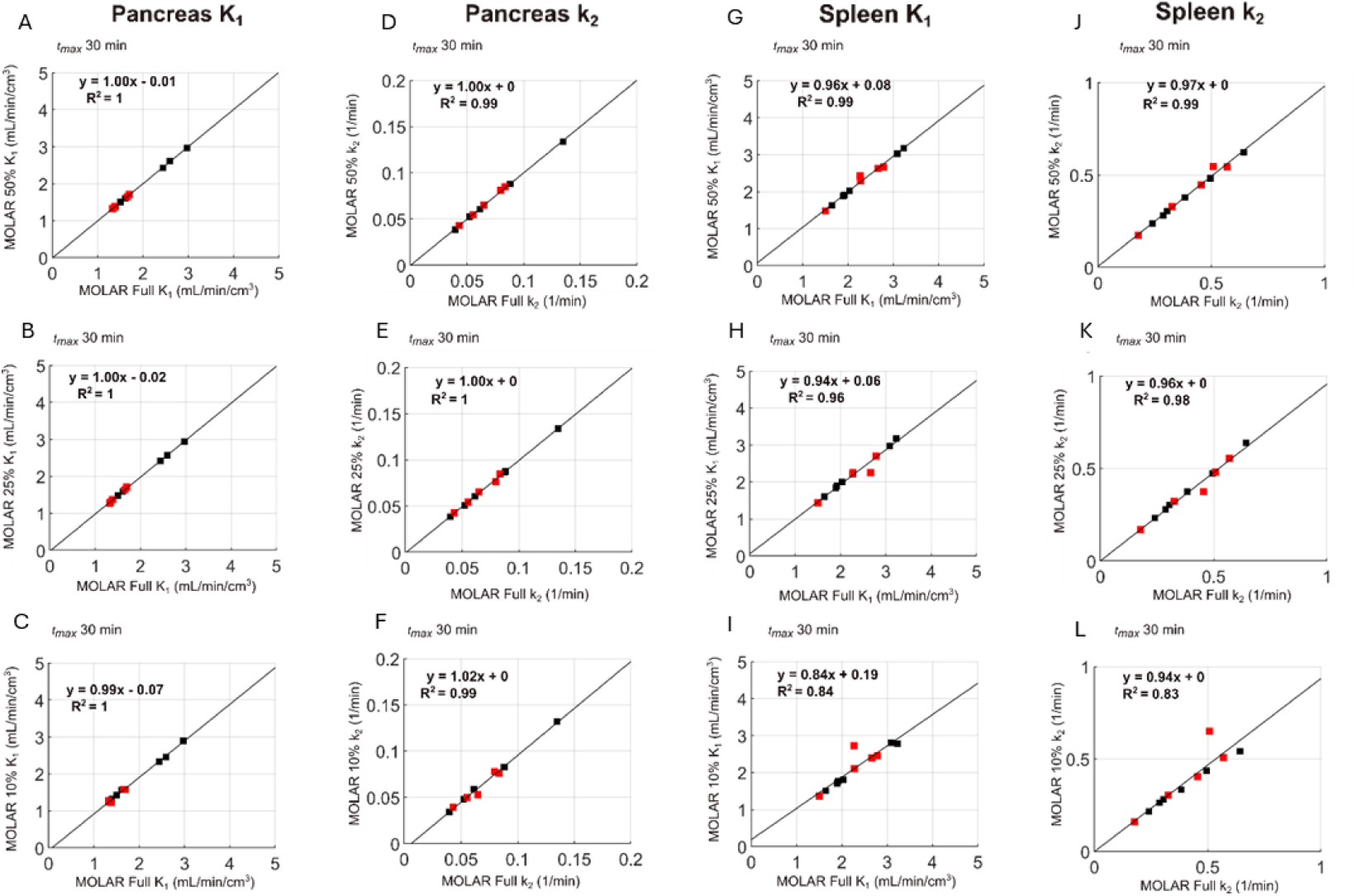
Scatter plots of volume of distribution (*V*_T_) estimates (1TC, *t*_max_=30) in healthy controls (black squares) and individuals with type 1 diabetes (red squares) for pancreas *K*_1_ (**A-C**), pancreas *k*_2_ (**D-F**), spleen *K*_1_ (**G-I**), and spleen *k*_2_ (**J-L**) between full-count and reduced count data. Black line represents linear regression fit.

**Supplemental Table 1.** Mean (Mean SE) of *K*_1_ and *k*2 estimates across all subjects for pancreas and spleen using 1TC model (*t*_max_=30 min) for full and reduced count data.

| Mean (Standard Error) | MOLAR Full Count | MOLAR 50% Count | MOLAR 25% Count | MOLAR 10% Count |
| --- | --- | --- | --- | --- |
| Pancreas $K_1$ | 1.81(0.08) | 1.81(0.08) | 1.79 (0.08) | 1.72(0.08) |
| Spleen $K_1$ | 2.30(0.18) | 2.29(0.18) | 2.21(0.18) | 2.13(0.16) |
| Pancreas $k_2$ | 0.07(0.01) | 0.07(0.01) | 0.07(0.01) | 0.07(0.01) |
| Spleen $k_2$ | 0.40(0.05) | 0.40(0.05) | 0.38(0.05) | 0.37(0.05) |

